# Cardiorespiratory Fitness Modifies the Relationship Between Arterial Stiffness and Cerebral Blood Flow Independent of Physical Activity

**DOI:** 10.1101/2025.03.03.25323254

**Authors:** B.M. Breidenbach, I. Driscoll, M.P. Glittenberg, A.J. Paulsen, S. Fernandes-Taylor, T. Naren, G.S. Roberts, T.L. Brach, M.M. Jarchow, L. E. Symanski, A.Y. Gaul, S.R. Lose, L.A. Rivera-Rivera, S.C. Johnson, S. Asthana, B.T. Christian, D.B. Cook, O. Wieben, O.C. Okonkwo

## Abstract

**INTRODUCTION:** Central arterial stiffness and cerebral blood flow (CBF) are inversely related. Poor cardiorespiratory fitness (CRF) and low physical activity (PA) are related to both higher arterial stiffness and lower CBF. The present study examined (i) whether CRF or PA moderate the relationship between arterial stiffness and CBF and (ii) whether the intensity or the type of PA need to be considered.

**METHODS:** Participants (N=78, Mean_AGE_=64.2±6.14, 72% female) from the Wisconsin Registry for Alzheimer’s Prevention and the Wisconsin Alzheimer’s Disease Research Center were categorized into low, average and high fitness groups based on maximal graded exercise treadmill test performance. PA was assessed using the CHAMPS questionnaire. Based on hours/week, participants were classified as meeting the recommended 2.5 hours of moderate intensity PA per week (PA Rec Met). Weekly hours of moderate and low intensity PA were calculated as activities of > 3 or < 3 metabolic equivalents, respectively. Activity type was categorized as exercise-, sports/leisure- and work-related. Arterial stiffness was measured as aortic pulse wave velocity (aoPWV) by 2D phase contrast MRI. CBF was assessed by 4D flow MRI in the internal carotid arteries (ICAs), cavernous ICAs, middle cerebral arteries (MCAs), and via two composite measures of total and global flow.

**RESULTS:** The association between aoPWV and CBF differed by fitness levels, with a negative relationship in the low fitness group and positive relationships in the average and high fitness groups (all *P*s<0.05). Significant moderating effects on the relationships between aoPWV and CBF were also observed for PA Rec Met (all *P*s<0.05), moderate intensity (*P*=0.05) and exercise-related (all *P*s<0.02) PA.

**DISCUSSION:** Average or high fitness, meeting the PA guidelines, and more specifically, moderate intensity and exercise-related PA seem to attenuate the negative relationship between aoPWV on CBF.

## 1 INTRODUCTION

Age-related arterial stiffening is associated with higher incidence of cardiovascular disease (CVD)^1^, cerebral small vessel disease^2^, unfavorable structural brain changes^3^, and cognitive impairment^4^. The literature also suggests a significant relationship between higher arterial stiffness and lower cerebral blood flow (CBF)^5^. A supporting theory posits that higher central arterial stiffness contributes to harmful pulsative blood flow to the cerebral microvasculature, triggering vasoconstriction as a protective compensatory mechanism, thereby increasing resistance and decreasing CBF^6,7^. CBF is essential for maintaining brain function and cognitive health, with cerebral hypoperfusion and cerebrovascular stiffening recognized as important risk factors contributing to cognitive decline and Alzheimer’s disease (AD)^6,8,9^. Individuals at higher risk for cognitive decline or dementia stand to benefit from strategies aimed at improving vascular health or lifestyle factors known to contribute to both CVD and AD.

Exercise and physical activity (PA) are known to improve outcomes related to cardiovascular and cerebrovascular health^10^, including arterial stiffness^11,12^ and cerebral hemodynamics^13,14^. Greater cardiorespiratory fitness (CRF), attained through regular aerobic exercise, seems to attenuate age-related central arterial stiffening^15,16^. Indeed, several exercise intervention studies have demonstrated beneficial changes in arterial stiffness with concurrent increases in maximal oxygen consumption (VO_2MAX_) indicating improvement in CRF^17^. Moreover, both high CRF and PA are associated with higher CBF^10,18–20^.

Although CRF is determined by a number of nonmodifiable factors such as age, sex, and genetic factors^21^, it is generally considered an objective indicator of habitual PA, with CRF and PA often used interchangeably^22^. While CRF is partly a consequence of regular PA, PA-related health benefits can occur independent of changes to CRF^21,23^. It is currently unknown whether each makes a unique contribution to CBF alterations related to central arterial stiffness. Given the beneficial relationship between CRF, PA, and vascular health (both cardiovascular and cerebrovascular), the present study examines whether CRF or PA moderate the relationship between arterial stiffness and CBF, and whether the intensity or the type of PA impact this relationship differentially.

## 2 MATERIALS AND METHODS

### 2.1 Participants

The sample consisted of 78 cognitively unimpaired adults (Mean_AGE_=64.2±6.14, 72% female) from the Wisconsin Registry for Alzheimer’s Prevention (WRAP)^24^ or the Wisconsin Alzheimer’s Disease Research Center (WADRC)^9^, both longitudinally followed cohorts enriched at enrollment for family history of dementia and *APOE* ε4 allele carriage^9,24^. Pariticpants for the present study were selected based on completion of a graded exercise treadmill test and cardiac and cranial MRI scans within 1 yr of each other. Socio-demographic and health characteristics are detailed in Table 1. All study procedures were approved by the University of Wisconsin Institutional Review Board and signed written consent was obtained from all participants.

**Table 1.** Participant Characteristics.

|  | **Total** | **Low**<br>**Fitness**<br>**(L)** | **Average**<br>**Fitness (A)** | **High**<br>**Fitness (H)** | **P** | **Post hoc** |
| --- | --- | --- | --- | --- | --- | --- |
|  | (N=78) | (N=14) | (N=20) | (N=44) |  |  |
| **<u>Demographic and health characteristics</u>** |  |  |  |  |  |  |
| **Age** *M (SD)* | 64.2 (6.14) | 64.9 (6.01) | 62.1 (7.07) | 64.9 (5.63) | 0.22 |  |
| **Female** *N (%)* | 56 (72) | 10 (71) | 14 (70) | 32 (73) | 0.1 |  |
| **Education**<br>(years; *M (SD)*) | 16.3 (2.21) | 15.0 (2.42) | 16.9 (1.87) | 16.5 (2.17) | **0.04** | A > L |
| **APOE ε4** *N (%)* | 38 (49) | 7 (50) | 7 (35) | 24 (55) | 0.35 |  |
| **Family Hx Dementia**<br>*N (%)* * | 39 (50) | 6 (43) | 8 (40) | 25 (57) | 0.18 |  |
| **WHR** *M (SD)* | 0.90(0.08) | 0.91 (0.06) | 0.92 (0.08) | 0.88 (0.08) | 0.2 |  |
| **Hypertension** *N (%)* | 18 (23) | 3 (21) | 8 (40) | 7 (16) | 0.1 |  |
| **aoPWV** (*m/s; M (SD)*) | 7.62 (3.57) | 7.86 (4.82) | 7.59 (3.06) | 7.56 (3.42) | 0.96 |  |
| **Elevated PWV**<br>(*>10m/s; N (%)*) | 13 (17) | 3 (21) | 4 (20) | 6 (14) | 0.71 |  |
| **VO<sub>2</sub>predicted**<br>(*ml/kg/min; M (SD)*) | 25.9 (6.13) | 18.9 (3.58) | 24.1 (3.90) | 29.0 (5.44) | **<0.001** | H > A > L |
| **<u>Physical Activity</u>** |  |  |  |  |  |  |
| **PA Rec Met**<br>(yes; *N (%)*) | 52 (67) | 7 (50) | 11 (55) | 34 (77) | 0.07 |  |
| **Low Intensity PA**<br>(*hrs/week; M (SD)*) | 7.94 (4.94) | 7.61 (4.97) | 7.53 (4.81) | 8.23 (5.08) | 0.84 |  |
| **Moderate Intensity PA**<br>(*hrs/week; M (SD)*) | 9.26 (7.01) | 7.34 (9.14) | 7.55 (6.79) | 10.7 (6.15) | 0.14 |  |
| **Exercise-Related PA**<br>(*hrs/week; M (SD)*) | 5.59 (4.74) | 3.18 (4.44) | 4.98 (4.96) | 6.64 (4.50) | **0.04** | H > A |
| **Leisure/Sports-Related PA**<br>(*hrs/week; M (SD)*) | 0.73 (1.94) | 0.45 (1.53) | 0.44 (1.05) | 0.10 (2.34) | 0.51 |  |
| **Work-Related PA**<br>(hrs/week; M (SD)) | 3.06 (3.63) | 3.71 (5.45) | 2.33 (2.93) | 3.19 (3.23) | 0.52 |  |
| **<u>Average CBF (mL/min; M (SD))</u>** |  |  |  |  |  |  |
| **Total Q** | 542 (99.1) | 522 (95.7) | 567 (84.0) | 538 (106) | 0.39 |  |
| **Global Q** | 126 (22.6) | 120 (21.4) | 131 (18.2) | 126 (24.6) | 0.34 |  |
| **ICA** | 211 (37.0) | 200 (34.3) | 225 (29.4) | 208 (39.6) | 0.12 |  |
| **Cav. ICA** | 220 (37.6) | 200 (31.7) | 236 (30.9) | 219 (39.6) | **0.02** | A > L |
| **MCA** | 131 (22.8) | 126 (20.9) | 137 (22.9) | 130 (23.3) | 0.32 |  |

\*Family Hx Dementia had 22% missingness, 28% negative, and 50% positive.
Post hoc: $H > A > L$ = high fitness mean is greater than means for average fitness and low fitness and average fitness mean is greater than low fitness mean; $H > A$ = high fitness mean is greater than average fitness mean; $A > L$ = average fitness mean is greater than low fitness mean.
**Abbreviations:** **APOE $\epsilon 4+$** = APOE $\epsilon 4$ allele carriage; **aoPWV** = aortic pulse wave velocity; **BA** = Basilar artery; **Cav. ICA** = cavernous ICA; **Exercise-Related PA** = exercise-related physical activity (hours/week); **Fitness** = Low, average, and high fitness groups were defined based on their calculated $VO_{2predicted}$ falling within the fitness percentile ranges for age and sex: $\leq 49\%$ , 50%-60%, and $\geq 70\%$ , respectively; **Global Q** = global flow; the average of seven cerebral arteries; **HTN** = hypertension; **ICA** = internal carotid arteries; **Low Intensity PA** = all physical activity less than 3.0 METs from the CHAMPS questionnaire; **MCA** = middle cerebral artery; **Moderate Intensity PA** = all physical activity greater than 3.0 METs from the CHAMPS questionnaire (hours/week); **M (SD)** = Mean (Standard Deviation); **PA Rec Met** = Physical Activity Recommendations of 150 minutes of moderate-intensity activity were met (yes/no); **Sports-Related PA** = sports/leisure related physical activity (hours/week); **Total Q** = total flow; the sum of the ICAs and the BA; **$VO_{2predicted}$** = predicted maximal oxygen consumption based on the oxygen uptake efficiency slope; **WHR** = waist to hip ratio; **Work-Related PA** = work related physical activity (hours/week).

### 2.2 Cardiorespiratory Fitness and Physical Activity

Participants performed a continuous graded exercise treadmill test (ETT; Trackmaster TMX428CP, Full Vision Inc., Newton, KS); full methods have been previously described^25^. Continuous measurements of oxygen uptake (VO_2_), carbon dioxide production (CO_2_), and minute ventilation (VE) were obtained using a metabolic cart and 2-way non-rebreathing valve (TrueOne 2400, ParvoMedics, Sandy, UT). As previously published, a predicted peak VO_2_ (VO_2predicted_; mL/kg/min) was calculated from metabolic data collected during the ETT based on the oxygen uptake efficiency slope (OUES) equation^26^ to determine CRF. For analyses purposes, participants were categorized into low (N=14), average (N=20), and high (N=44) fitness groups based on age and sex specific reference standards for VO_2_ established through normative data from the “FRIEND” (Fitness Registry and the Importance of Exercise) National Data Base^27^; with low fitness falling below the 49^th^ percentile, average fitness between the 50-69^th^ percentile, and high fitness at or above the 70^th^ percentile.

At the time of ETT assessment, participants also completed the CHAMPS physical activity questionnaire (Community Health Activities Model Program for Seniors). CHAMPS is designed for older adults and has an established reliability, sensitivity, and construct validity^28^. Of the total hours/week of PA (Total PA), moderate and low intensity PA were respectively defined as hours/week of ≥ 3 or < 3 metabolic equivalents (METs) activities. Those with ≥ 2.5 hours of moderate intensity PA per week were considered to have met the PA recommendations (PA Rec Met) as set by the World Health Organization (WHO)^29^. Of 52 participants who were classified as physically active determined by moderate intensity activities, 25 also completed some level of vigorous intensity activity (≥6 METs). However, we opted to combine them under the moderate intensity grouping for analyses purposes because the low variation in vigorous activity rendered subgroups underpowered when modeling vigorous PA as a separate group. PA type was grouped either as exercise-, sports/leisure- and work-related based on activity descriptions by WHO^29^.

Instructions and further details on scoring PA from the questionnaire for type and intensity are detailed in Figure 1.

**Figure 1.**
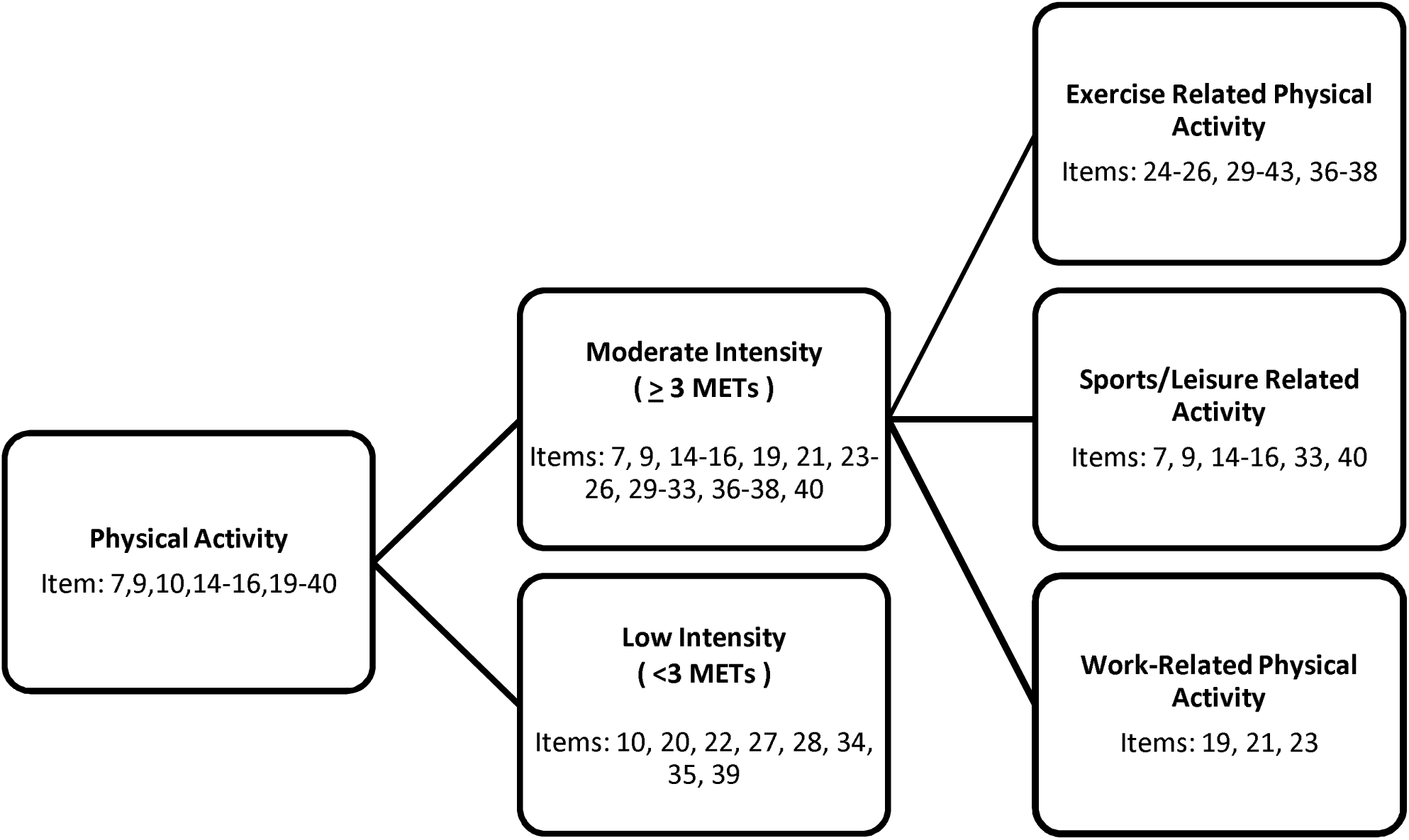
**Physical Activity Scoring Instructions** CHAMPS Questionnaire items for each category are reflected in the figure. The intensity for each activity (METs) as well as total physical activity (hours/week) was calculated as the sum of all activities according to the original reference^28^. Meeting the Physical Activity Recommendations (PA Rec Met) was calculated as achieving ≥ 2.5 hours (150 minutes) of moderate intensity activity in a week. Moderate-intensity PA (hours/week) was defined as any PA ≥ 3 METs, while low-intensity PA (hours/week) was defined as any PA < 3 METs^28^. Exercise, leisure/sports, and work related activities were classified as such based on the World Health Organization’s “Global Recommendations on Physical Activity for Health”^29^. Exercise-related PA was defined as activities traditionally aimed at increasing a component of physical fitness like cycling, swimming, running, walking, or strength training. Leisure/sports-related activities were defined as leisure or recreational related sporting activities, like dancing, golf, basketball, soccer, racquetball, and gentle swimming and work-related activities were defined as tasks performed to maintain and manage a home or as part of an occupation.

### 2.3 Cardiovascular Magnetic Resonance (CMR) Imaging for Arterial Stiffness

Central arterial stiffness was assessed with aortic pulse wave velocity (aoPWV; meters/second) measurements obtained with magnetic resonance imaging (MRI). MRI data were acquired as previously described in detail^30^ on a clinical 3T scanners (Discovery MR 750 and Signa Premier, GE Healthcare, Waukesha, WI) using an 8-channel torso coil. An ungated, breath-hold, 2D multi-slice balanced steady-state free-precession (bSSFP) product sequence generated anatomical images to guide plane placement of the flow measurement planes and for aortic centerline tracing and subsequent distance measurements between the planes. A 2D radially undersampled, velocity-sensitive MRI sequence with retrospective cardiac and respiratory gating was used to enable free breathing acquisitions^30^. Flow data were acquired in two axial planes: one over the aortic arch including the ascending and descending aorta and one in the abdominal aorta above the renal arteries allowing for three flow measurements. Photoplethysmography (PPG) data were recorded using a pulse oximeter on the index finger and respiratory motion was recorded using bellows on the chest. Time-resolved and time-averaged velocity and magnitude datasets were reconstructed from data during expiration with a respiratory acceptance window of 50%. A customized analysis package with graphical user interface (GUI) was developed for PWV calculations using MATLAB (The Mathworks, Natwick, MA) and is publicly available (https://github.com/tnaren97/PWV_2DPC). The processing pipeline includes computation of centerlines and flow plane distances from the anatomical bSSFP images and determines time shifts between the flow waveforms using the Time-to-Point (TTP) PWV method^31^. Flow waveforms were smoothed with a Gaussian filter (width=7 pixels) and linearly temporally interpolated (20x).

### 2.4 4D Flow Magnetic Resonance Imaging and Analysis

As previously published^9^, a cranial MRI scan acquisition was performed on one of the two GE Healthcare 3T clinical systems: Discovery MR750 with a 32-channel head coil or Signa Premier with a 48-channel coil. Cerebral artery flow was assessed using a radial 4D Flow MRI sequence^32,33^.

Automatic vessel segmentation and flow quantification were performed in a customized post-processing tool (MATLAB, Mathworks, Natick, MA). Bilateral cycle-averaged volumetric flow rates (mL/min) were obtained in the distal cervical internal carotid arteries (ICAs), cavernous ICAs (Cav. ICAs), middle cerebral arteries (MCAs), vertebral arteries (VAs), anterior cerebral arteries (ACAs), posterior cerebral arteries (PCAs) and the basilar artery (BA). Total cerebral inflow (Total Q) reflected the sum of the left and right ICAs and BA. A composite score for global flow (Global Q) was created by averaging flows from the bilateral ICAs, Cav. ICAs, MCAs, VAs, ACAs, PCAs, and the BA. CBF outcomes utilized for analyses purposes were CBF in the ICA, Cav. ICA, MCA, Total Q and Global Q; and were selected based on the existent literature^18,34^.

### 2.5 Covariate Definitions

All models were adjusted for age, sex, *APOE* ε*4* allele presence, waist to hip ratio, and hypertension. Covariates were determined *a priori* based on the existing literature^5,35–37^. Waist to hip ratio (WHR) was calculated as waist circumference (cm) divided by hip circumference (cm) and was selected over body mass index to better represent central adiposity/obesity. Hypertension was defined as self-report or based on medication use. Being a carrier of at least one *APOE* ε4 allele (*APOE*4+) was treated as a categorical variable (positive/negative).

### 2.6 Statistical Analyses

Analyses were performed using R Statistical Software 4.4.0 (www.r-project.org). Data visualization was conducted using the ggplot2 package. For all regression models, assumptions of linearity, normality of residuals, homoscedasticity, and absence of multicollinearity were checked using regression diagnostics obtained from the Performance package in R. Sample characteristics were evaluated using one-way analysis of variance (ANOVA) for continuous or χ^2^ test for categorical measures. Tukey HSD post hoc tests were conducted to compare participant characteristics between low, average, and high fitness groups.

Covariate adjusted, linear regression models with CBF outcomes (Total Q, Global Q and mean bilateral blood flow in the ICA, Cav. ICA, and MCA) were used to address the aims.. Analyses were first performed with CRF as the predictor while covarying for PA Rec Met to investigate the moderating effect of CRF (low, average, high) on the relationship between aoPWV and CBF; the interaction between CRF and aoPWV was the term of interest. Then the analyses were repeated with PA Rec Met as the predictor while covarying for CRF (VO_2predicted)_) to parse out the contribution of PA to overall CRF, in moderating the relationship between aoPWV and CBF; the interaction between PA Rec Met (yes/no) and aoPWV was the term of interest. We further examined whether the moderating effect of PA on the relationship between aoPWV and CBF was dependent on PA intensity (moderate or low) and type (exercise, sports/leisure, and work-related).

## 3 RESULTS

### 3.1 Sample Characteristics

Table 1 details sociodemographic and health characteristics of study participants. The sample was predominantly white (94%) and female (72%), with an average age of 64.2 ± 6.14. The sample was enriched for AD risk at enrollment, with 49% carrying at least one *APOE* ε*4* allele and 50% having parental history of dementia. Overall, the sample had low to moderate prevalence of hypertension (23%), aoPWV ranged from 3.02 to 23.2 m/s and 16% of the sample had what would clinically (>10 m/s) be considered elevated aoPWV^38^.

Based on reference standards for VO_2predicted_, 18% of the sample was classified as low, 26% as average, and 56% as highly fit. There were no significant differences in age (*P*=0.22), sex (*P*=0.1), hypertension (*P*=0.1), *APOE*4+ status (*P*=0.35), or aoPWV (*P* =0.96) among fitness groups. The low fitness group was significantly less educated compared to the average (*P*=0.04), but not the high (*P* =0.07) fitness group. No group differences were found in hours/week engaged in low (*P*=0.84) or moderate (*P*=0.14) intensity PA. There were also no significant group differences in hours/week engaged in leisure/sports (*P*=0.51) or work-related (*P*=0.52) PA. The high fitness group averaged significantly more exercise-related activity compared to the low fitness group (*P*=0.04); no differences were observed between average and high (*P*=0.4) or between average and low (*P*=0.5) fitness groups. The low fitness group had a significantly lower CBF only in the Cav. ICA (*P*=0.02) compared to the average fitness group.

### 3.2 CRF- and PA-related alterations in cerebral blood flow

Results for the joint effect of CRF and aoPWV on CBF, adjusted for PA Rec Met, are presented in Table 2 and Supplemental Figure 1. There were significant positive interactions between the high fitness group and aoPWV for all CBF outcomes [Total Q (*P*=0.02), Global Q (*P*=0.04), ICA (*P*=0.01), Cav. ICA (*P*=0.03), MCA (*P*=0.02)], as well as between the average fitness group and aoPWV for Total Q (*P*=0.02) and in the ICA (*P*=0.01), Cav. ICA (*P*=0.02), and MCA (*P*=0.01), where for each 1 unit increase in aoPWV, those in the low fitness group had decreased CBF while those in the high and average fitness group increased CBF. In the low fitness group, higher aoPWV was related to lower CBF [Total Q (*P*=0.04); Global Q (*P*=0.05); ICA (*P*=0.03); Cav. ICA (P=0.06); MCA (*P*=0.05)]. Age was negatively associated with CBF for Global Q (*P*=0.01), and in the Cav. ICA (*P*=0.03) and MCA (*P*=0.01). Hypertension was positively associated with CBF for Total Q (*P*=0.03), and in the ICA (*P*=0.02), and MCA (*P*=0.03).

**Table 2.** Associations between arterial stiffness and cardiorespiratory fitness on cerebral blood flow, adjusting for physical activity.

| Outcome | Exposure | Estimate | Std.Error | P |
| --- | --- | --- | --- | --- |
| Total Q | Age | -3.73 | 2.01 | 0.07 |
|  | WHR | -321.65 | 204.41 | 0.12 |
|  | **HTN** | 64.13 | 28.54 | **0.03** |
|  | **aoPWV** | -12.14 | 5.64 | **0.04** |
|  | PA Rec Met (yes) | -35.25 | 24.61 | 0.16 |
|  | **aoPWV \* average fitness** | 22.63 | 9.61 | **0.02** |
|  | **aoPWV \* high fitness** | 17.09 | 7.16 | **0.02** |
| Global Q | **Age** | -1.17 | 0.46 | **0.01** |
|  | WHR | -52.49 | 47.01 | 0.27 |
|  | HTN | 12.41 | 6.56 | 0.06 |
|  | **aoPWV** | -2.59 | 1.30 | **0.05** |
|  | PA Rec Met (yes) | -6.93 | 5.66 | 0.23 |
|  | aoPWV * average fitness | 3.97 | 2.21 | 0.08 |
|  | **aoPWV \* high fitness** | 3.54 | 1.65 | **0.04** |
| ICA | Age | -1.02 | 0.73 | 0.17 |
|  | WHR | -134.65 | 73.82 | 0.07 |
|  | **HTN** | 24.81 | 10.31 | **0.02** |
|  | **aoPWV** | -4.55 | 2.04 | **0.03** |
|  | PA Rec Met (yes) | -17.01 | 8.89 | 0.06 |
|  | **aoPWV \* average fitness** | 9.53 | 3.47 | **0.01** |
|  | **aoPWV \* high fitness** | 6.85 | 2.59 | **0.01** |
| Cav. ICA | **Age** | -1.57 | 0.72 | **0.03** |
|  | WHR | -138.61 | 73.31 | 0.06 |
|  | HTN | 18.48 | 10.24 | 0.08 |
|  | aoPWV | -3.86 | 2.02 | 0.06 |
|  | PA Rec Met (yes) | -17.37 | 8.83 | 0.06 |
|  | **aoPWV \* average fitness** | 8.37 | 3.45 | **0.02** |
|  | **aoPWV \* high fitness** | 5.82 | 2.57 | **0.03** |
| MCA | **Age** | -1.21 | 0.45 | **0.01** |
|  | WHR | -83.43 | 45.67 | 0.07 |
|  | **HTN** | 14.44 | 6.38 | **0.03** |
|  | **aoPWV** | -2.52 | 1.26 | **0.05** |
|  | PA Rec Met (yes) | -4.83 | 5.50 | 0.38 |
|  | **aoPWV \* average fitness** | 5.41 | 2.15 | **0.01** |
|  | **aoPWV \* high fitness** | 3.87 | 1.60 | **0.02** |

Covariates: PA Rec Met, age, sex, WHR, HTN, and APOE $\epsilon 4+$ . Only significant covariates and the term(s) of interest (interaction) were included in the table.
Abbreviations: **APOE $\epsilon 4+$** = APOE $\epsilon 4$ allele carriage; **aoPWV** = aortic pulse wave velocity; **BA** = Basilar artery; **Cav. ICA** = cavernous ICA; **Fitness** = Low, average, and high fitness groups were defined based on their calculated $VO_{2predicted}$ falling within the fitness percentile ranges for age and sex: $\leq 49\%$ , 50%-60%, and $\geq 70\%$ , respectively; **Global Q** = global flow; the average of seven cerebral arteries; **HTN** = hypertension; **ICA** = internal carotid arteries; **MCA** = middle cerebral artery; **PA Rec Met** = Physical Activity Recommendations of 150 minutes of moderate-intensity activity were met (yes/no); **Total Q** = total flow; the sum of the ICAs and the BA; **$VO_{2predicted}$** = predicted maximal oxygen consumption based on the oxygen uptake efficiency slope; **WHR** = waist to hip ratio.

Results for the joint effect of PA Rec Met and aoPWV on CBF, adjusted for CRF, are presented in Table 3 and Supplemental Figure 2. There were significant positive interactions between PA Rec Met and aoPWV on CBF for Total Q (*P*=0.03), Global Q (*P*=0.05) and in the MCA (*P*=0.03), such that the adverse relationship between aoPWV and CBF was attenuated in those meeting PA recommendations compared to those who did not. Age (*P*=0.03) and WHR (*P*= 0.05) were negatively associated with CBF in the MCA.

**Table 3.**
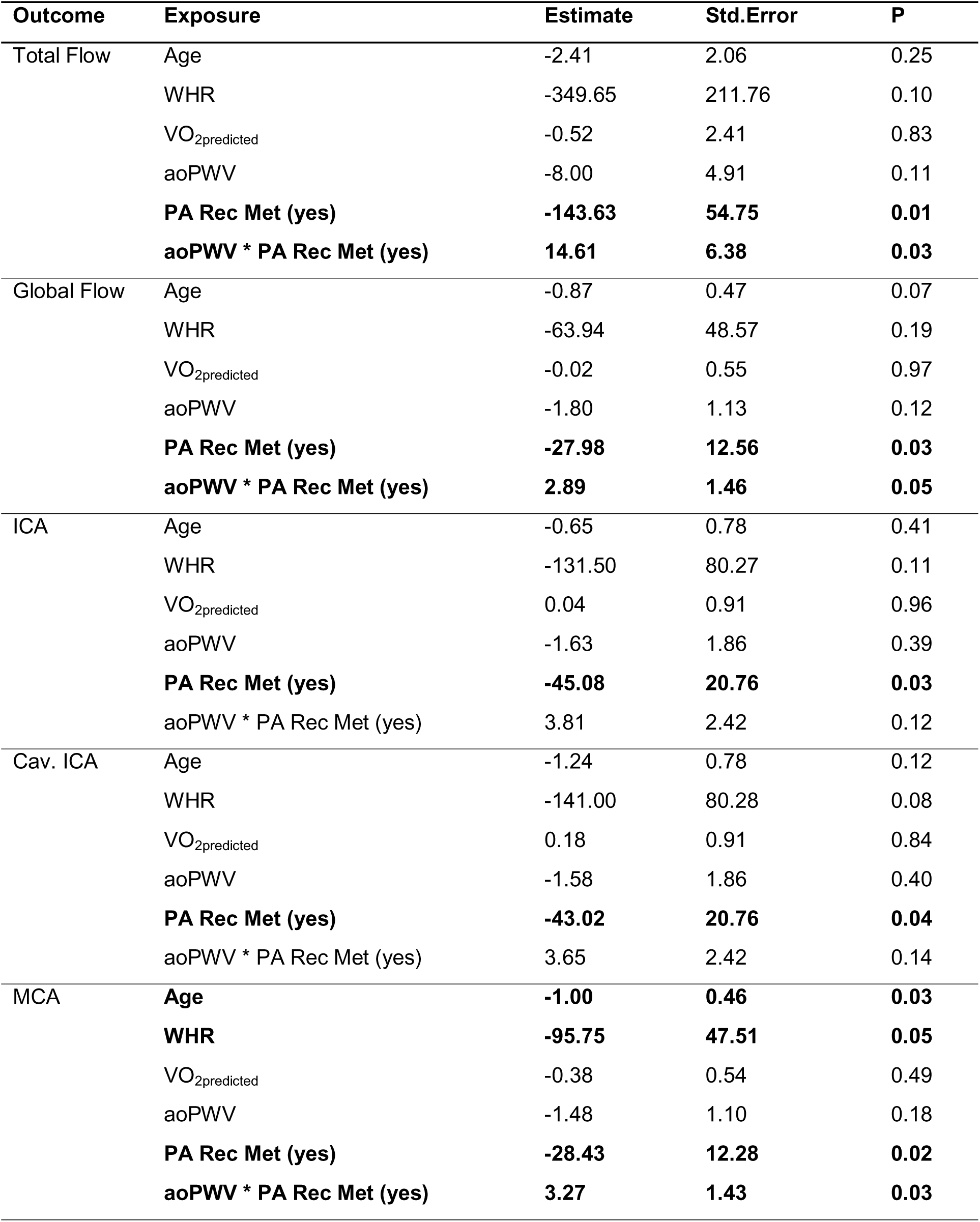
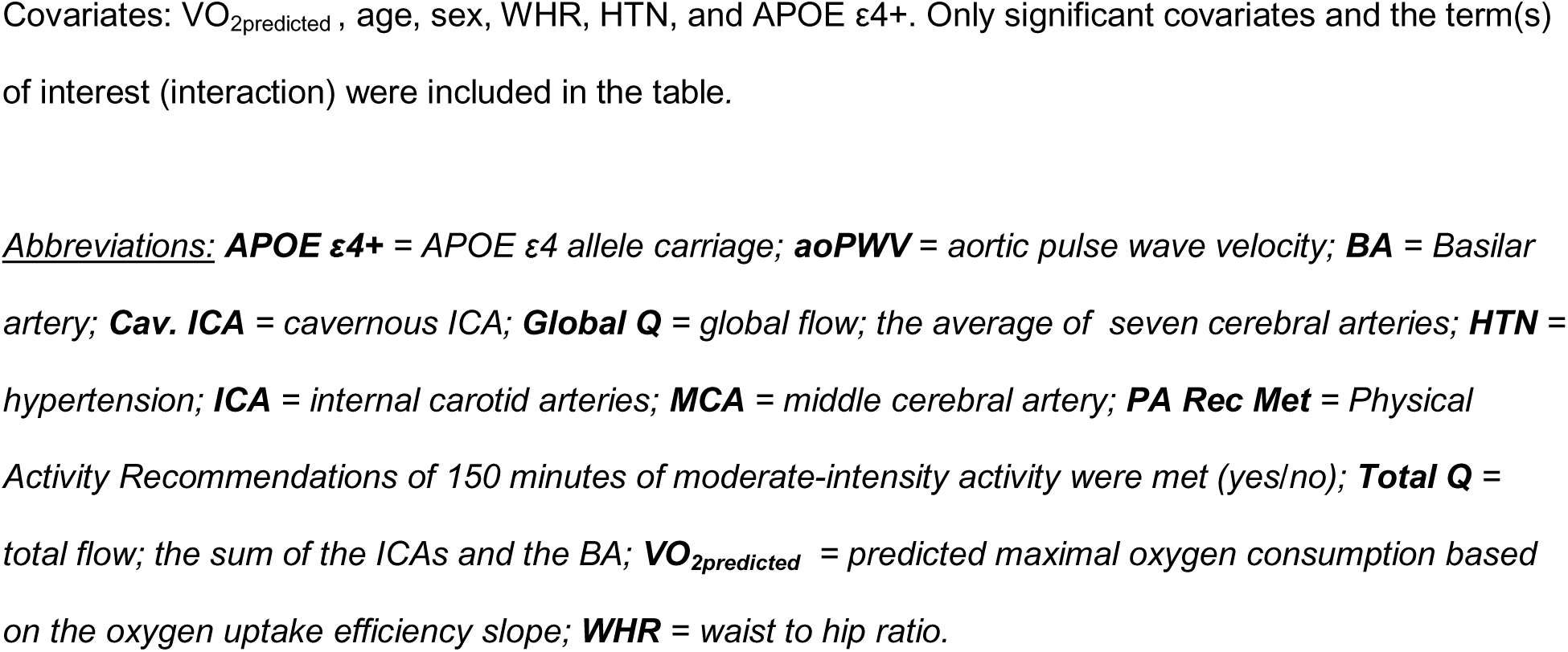
Associations between arterial stiffness and physical activity on cerebral blood flow, adjusting for cardiorespiratory fitness.

| Outcome | Exposure | Estimate | Std.Error | P |
| --- | --- | --- | --- | --- |
| Total Flow | Age | -2.41 | 2.06 | 0.25 |
|  | WHR | -349.65 | 211.76 | 0.10 |
|  | VO<sub>2</sub>predicted | -0.52 | 2.41 | 0.83 |
|  | aoPWV | -8.00 | 4.91 | 0.11 |
|  | **PA Rec Met (yes)** | **-143.63** | **54.75** | **0.01** |
|  | **aoPWV \* PA Rec Met (yes)** | **14.61** | **6.38** | **0.03** |
| Global Flow | Age | -0.87 | 0.47 | 0.07 |
|  | WHR | -63.94 | 48.57 | 0.19 |
|  | VO<sub>2</sub>predicted | -0.02 | 0.55 | 0.97 |
|  | aoPWV | -1.80 | 1.13 | 0.12 |
|  | **PA Rec Met (yes)** | **-27.98** | **12.56** | **0.03** |
|  | **aoPWV \* PA Rec Met (yes)** | **2.89** | **1.46** | **0.05** |
| ICA | Age | -0.65 | 0.78 | 0.41 |
|  | WHR | -131.50 | 80.27 | 0.11 |
|  | VO<sub>2</sub>predicted | 0.04 | 0.91 | 0.96 |
|  | aoPWV | -1.63 | 1.86 | 0.39 |
|  | **PA Rec Met (yes)** | **-45.08** | **20.76** | **0.03** |
|  | **aoPWV \* PA Rec Met (yes)** | **3.81** | **2.42** | **0.12** |
| Cav. ICA | Age | -1.24 | 0.78 | 0.12 |
|  | WHR | -141.00 | 80.28 | 0.08 |
|  | VO<sub>2</sub>predicted | 0.18 | 0.91 | 0.84 |
|  | aoPWV | -1.58 | 1.86 | 0.40 |
|  | **PA Rec Met (yes)** | **-43.02** | **20.76** | **0.04** |
|  | **aoPWV \* PA Rec Met (yes)** | **3.65** | **2.42** | **0.14** |
| MCA | **Age** | **-1.00** | **0.46** | **0.03** |
|  | **WHR** | **-95.75** | **47.51** | **0.05** |
|  | VO<sub>2</sub>predicted | -0.38 | 0.54 | 0.49 |
|  | aoPWV | -1.48 | 1.10 | 0.18 |
|  | **PA Rec Met (yes)** | **-28.43** | **12.28** | **0.02** |
|  | **aoPWV \* PA Rec Met (yes)** | **3.27** | **1.43** | **0.03** |

Covariates: $VO_{2\text{predicted}}$ , age, sex, WHR, HTN, and APOE $\epsilon 4+$ . Only significant covariates and the term(s) of interest (interaction) were included in the table.
Abbreviations: **APOE $\epsilon 4+$** = APOE $\epsilon 4$ allele carriage; **aoPWV** = aortic pulse wave velocity; **BA** = Basilar artery; **Cav. ICA** = cavernous ICA; **Global Q** = global flow; the average of seven cerebral arteries; **HTN** = hypertension; **ICA** = internal carotid arteries; **MCA** = middle cerebral artery; **PA Rec Met** = Physical Activity Recommendations of 150 minutes of moderate-intensity activity were met (yes/no); **Total Q** = total flow; the sum of the ICAs and the BA; **$VO_{2\text{predicted}}$** = predicted maximal oxygen consumption based on the oxygen uptake efficiency slope; **WHR** = waist to hip ratio.

### 3.3 Alterations in cerebral blood flow based on PA intensity and type

Results for PA intensity are presented in Table 4 and Supplementary Table 1. An investigation into the intensity of PA, categorized as moderate or low revealed that low-intensity PA did not significantly modify any of the relationships between aoPWV and CBF (all *P*s≥0.30; sTable 1). A significant positive interaction between moderate intensity PA and aoPWV was observed for Global Q (*P*=0.05; Table 4). Hypertension was also positively associated with CBF (Total Q: *P=0.03*; ICA: *P*=0.02; MCA: *P*=0.03) whereas, age was negatively associated with CBF (Global Q: *P*=0.03; Cav. ICA: *P*=0.04; MCA: *P*=0.02).

**Table 4.**
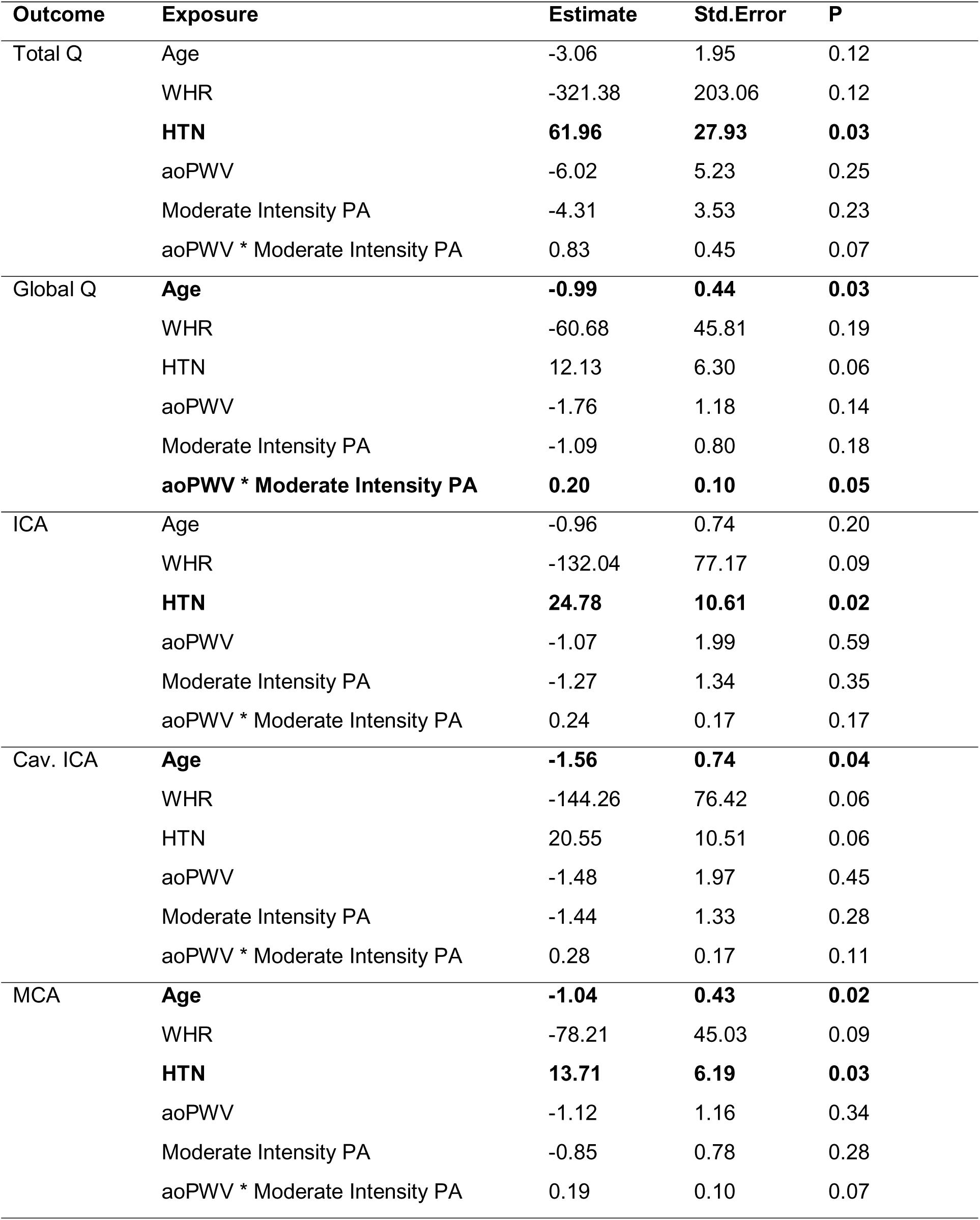
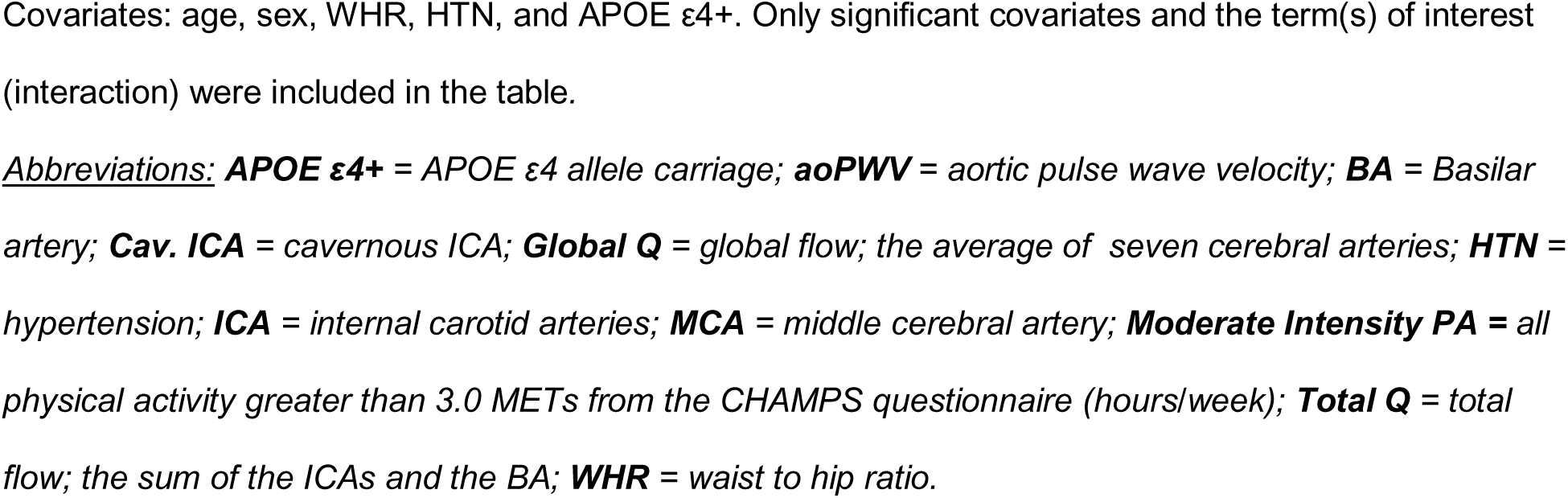
Associations between arterial stiffness and moderate intensity physical activity on cerebral.

| Outcome | Exposure | Estimate | Std.Error | P |
| --- | --- | --- | --- | --- |
| Total Q | Age | -3.06 | 1.95 | 0.12 |
|  | WHR | -321.38 | 203.06 | 0.12 |
|  | **HTN** | **61.96** | **27.93** | **0.03** |
|  | aoPWV | -6.02 | 5.23 | 0.25 |
|  | Moderate Intensity PA | -4.31 | 3.53 | 0.23 |
|  | aoPWV * Moderate Intensity PA | 0.83 | 0.45 | 0.07 |
| Global Q | **Age** | **-0.99** | **0.44** | **0.03** |
|  | WHR | -60.68 | 45.81 | 0.19 |
|  | HTN | 12.13 | 6.30 | 0.06 |
|  | aoPWV | -1.76 | 1.18 | 0.14 |
|  | Moderate Intensity PA | -1.09 | 0.80 | 0.18 |
|  | **aoPWV \* Moderate Intensity PA** | **0.20** | **0.10** | **0.05** |
| ICA | Age | -0.96 | 0.74 | 0.20 |
|  | WHR | -132.04 | 77.17 | 0.09 |
|  | **HTN** | **24.78** | **10.61** | **0.02** |
|  | aoPWV | -1.07 | 1.99 | 0.59 |
|  | Moderate Intensity PA | -1.27 | 1.34 | 0.35 |
|  | aoPWV * Moderate Intensity PA | 0.24 | 0.17 | 0.17 |
| Cav. ICA | **Age** | **-1.56** | **0.74** | **0.04** |
|  | WHR | -144.26 | 76.42 | 0.06 |
|  | HTN | 20.55 | 10.51 | 0.06 |
|  | aoPWV | -1.48 | 1.97 | 0.45 |
|  | Moderate Intensity PA | -1.44 | 1.33 | 0.28 |
|  | aoPWV * Moderate Intensity PA | 0.28 | 0.17 | 0.11 |
| MCA | **Age** | **-1.04** | **0.43** | **0.02** |
|  | WHR | -78.21 | 45.03 | 0.09 |
|  | **HTN** | **13.71** | **6.19** | **0.03** |
|  | aoPWV | -1.12 | 1.16 | 0.34 |
|  | Moderate Intensity PA | -0.85 | 0.78 | 0.28 |
|  | aoPWV * Moderate Intensity PA | 0.19 | 0.10 | 0.07 |

Covariates: age, sex, WHR, HTN, and APOE $\epsilon 4+$ . Only significant covariates and the term(s) of interest (interaction) were included in the table.
Abbreviations: **APOE $\epsilon 4+$** = APOE $\epsilon 4$ allele carriage; **aoPWV** = aortic pulse wave velocity; **BA** = Basilar artery; **Cav. ICA** = cavernous ICA; **Global Q** = global flow; the average of seven cerebral arteries; **HTN** = hypertension; **ICA** = internal carotid arteries; **MCA** = middle cerebral artery; **Moderate Intensity PA** = all physical activity greater than 3.0 METs from the CHAMPS questionnaire (hours/week); **Total Q** = total flow; the sum of the ICAs and the BA; **WHR** = waist to hip ratio.

We subsequently investigated whether different types of moderate intensity PA (exercise, sports/leisure, and work-related activities) modify the aoPWV-CBF relationship. Results are displayed in Table 5 and Supplementary Tables 2-3. There were significant positive interactions between exercise-related PA and aoPWV on CBF (Table 5) for Total Q (*P*=0.02), Global Q (*P*=0.02), and in the MCA (*P*=0.02). Hypertension was positively associated with CBF (ICA: *P*= 0.05), while age was negatively associated with CBF (Global Q: *P=0.04*; MCA: *P*=0.04). Neither sports/leisure (all *P*s>0.12; sTable 2) nor work-related PA (all *P*s > 0.1; sTable 3) modified the effect of aoPWV on CBF. A positive main effect was found between sports/leisure-related PA and CBF for Total Q (*P*=0.04), Global Q (*P*=0.03), and in the MCA (*P*=0.04), indicating higher CBF with higher amounts of sports/leisure-related PA achieved. Hypertension was also positively associated with CBF in the ICA (*P*=0.04), while age was negatively associated with CBF for Global Q (*P*=0.01), and in the Cav. ICA (*P*=0.02) and MCA (*P*=0.01).

**Table 5.**
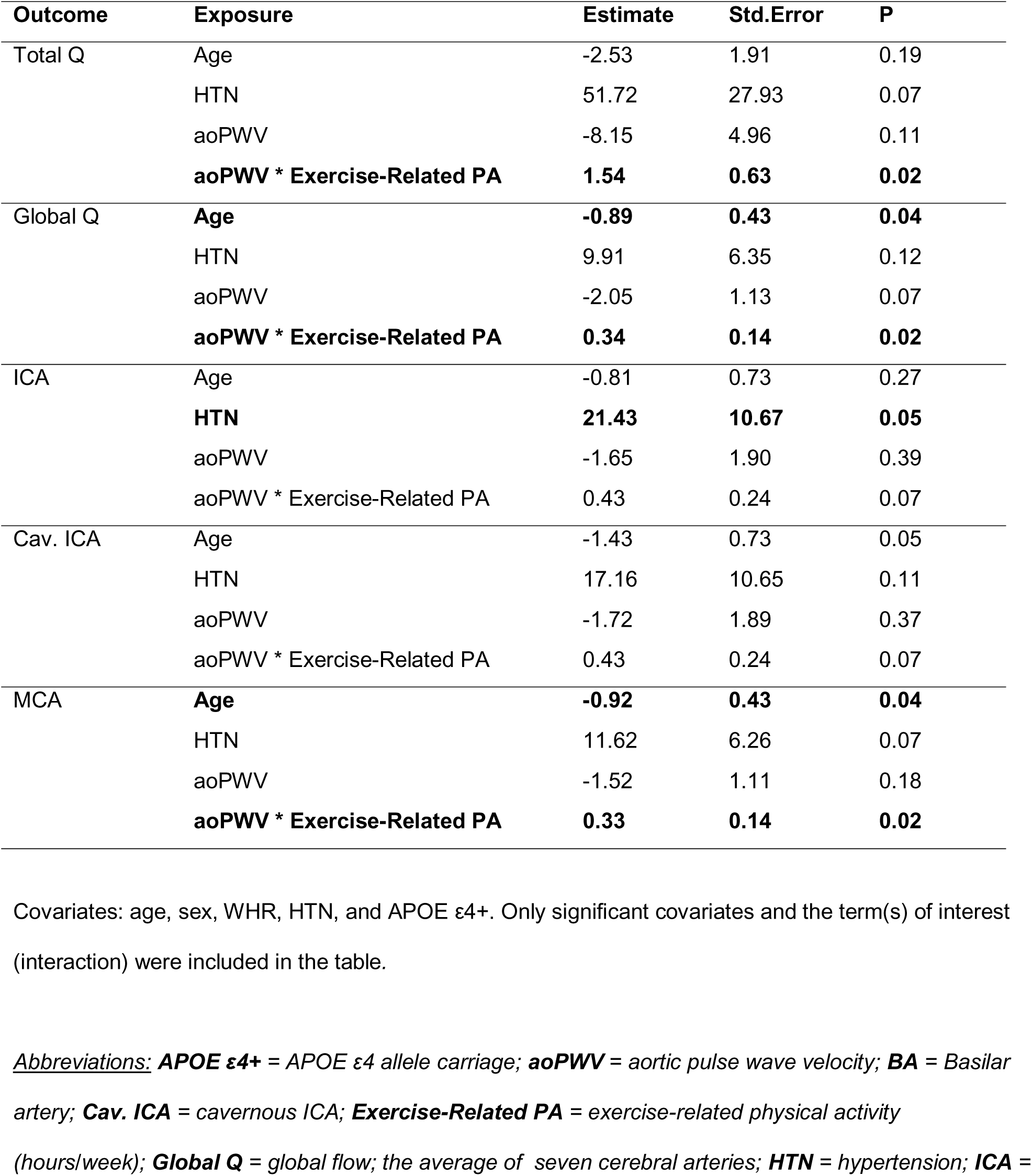
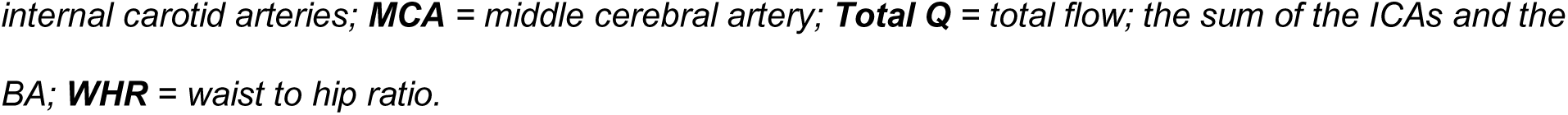
Associations between arterial stiffness and exercise-related physical activity on cerebral blood.

| Outcome | Exposure | Estimate | Std.Error | P |
| --- | --- | --- | --- | --- |
| Total Q | Age | -2.53 | 1.91 | 0.19 |
|  | HTN | 51.72 | 27.93 | 0.07 |
|  | aoPWV | -8.15 | 4.96 | 0.11 |
|  | **aoPWV \* Exercise-Related PA** | **1.54** | **0.63** | **0.02** |
| Global Q | **Age** | **-0.89** | **0.43** | **0.04** |
|  | HTN | 9.91 | 6.35 | 0.12 |
|  | aoPWV | -2.05 | 1.13 | 0.07 |
|  | **aoPWV \* Exercise-Related PA** | **0.34** | **0.14** | **0.02** |
| ICA | Age | -0.81 | 0.73 | 0.27 |
|  | **HTN** | **21.43** | **10.67** | **0.05** |
|  | aoPWV | -1.65 | 1.90 | 0.39 |
|  | aoPWV * Exercise-Related PA | 0.43 | 0.24 | 0.07 |
| Cav. ICA | Age | -1.43 | 0.73 | 0.05 |
|  | HTN | 17.16 | 10.65 | 0.11 |
|  | aoPWV | -1.72 | 1.89 | 0.37 |
|  | aoPWV * Exercise-Related PA | 0.43 | 0.24 | 0.07 |
| MCA | **Age** | **-0.92** | **0.43** | **0.04** |
|  | HTN | 11.62 | 6.26 | 0.07 |
|  | aoPWV | -1.52 | 1.11 | 0.18 |
|  | **aoPWV \* Exercise-Related PA** | **0.33** | **0.14** | **0.02** |

Covariates: age, sex, WHR, HTN, and APOE $\epsilon 4+$ . Only significant covariates and the term(s) of interest (interaction) were included in the table.
**Abbreviations:** **APOE $\epsilon 4+$** = APOE $\epsilon 4$ allele carriage; **aoPWV** = aortic pulse wave velocity; **BA** = Basilar artery; **Cav. ICA** = cavernous ICA; **Exercise-Related PA** = exercise-related physical activity
(hours/week); **Global Q** = global flow; the average of seven cerebral arteries; **HTN** = hypertension; **ICA** =
internal carotid arteries; **MCA** = middle cerebral artery; **Total Q** = total flow; the sum of the ICAs and the BA; **WHR** = waist to hip ratio.

## 4 DISCUSSION

The overreaching goal of this study was to characterize the relationships between CRF or PA, aoPWV, and CBF in a cognitively unimpaired, late middle-aged cohort enriched for AD risk. Our findings suggest that having average or high CRF or meeting PA recommendations attenuates the adverse relationship between arterial stiffness and CBF. Importantly, the beneficial impact of average or high CRF on the aoPWV–CBF relationship remained significant even after adjusting for PA status, and *vice versa*, suggesting that CRF and PA account for unique variance in outcomes (i.e., they are not completely overlapping constructs). Further investigations into the intensity and type of PA suggest that the PA-related findings were likely driven by individuals with greater CRF, as only moderate intensity and exercise-related PA modified the aoPWV-CBF relationship, both of which are activities with high potential to increase CRF (compared to low intensity or sports/leisure related PA).

In contrast to well established negative relationships between hypertension and CBF^39,40^, hypertension was related to better CBF in our sample. Further investigation revealed that 12 of 18 participants with hypertension were currently receiving treatment [angiotensin-converting enzyme inhibitors (ACE, n=3), angiotensin receptor blockers (ARBs, n=1), calcium channel blockers (CCBs, n=4), hydrochlorothiazide diuretic (HCTZs, n=1) or a combination of beta-blockers with one of the previously mentioned treatments (n=3)]. Adherence to and the length of treatment are unknown. Of the 12 hypertensive participants receiving treatment, 11 were on vasodilators (ACE, ARBs, CCBs) known to increase blood flow in the brain^41^, which may have at least partly contributed to this unexpected finding.

In comparison to the existing literature, where CRF and PA are investigated separately^18,19,42^, we were able to assess both simultaneously and in the same sample for the present study. Moreover, the inclusion of fitness categories and the differentiation of PA scores were an important feature of our study that allowed greater insight into the type and intensity of PA that may moderate relationships between arterial stiffness and CBF. Published literature indicates that higher CRF and PA are associated with lower arterial stiffness^15,17^, and that higher CRF and PA are also independently associated with greater CBF^14,20^ and better cognitive performance^25^. Moreover, longitudinal improvements in CRF (VO_2peak_) coincide with reductions in arterial stiffness and CBF increases in cognitively unimpaired^18^ and impaired^19^ individuals. We observed a non-significant trend of moderate fitness having the greatest attenuation of the adverse aoPWV–CBF relationship (Supplementary Figure 1). Interestingly, Liu et al., also reported that moderate intensity continuous training improved CBF and executive function more effectively compared to high intensity interval training, albeit in young adults^13^.

We did not observe significant differences in aoPWV across the three fitness groups (Table 1), which contrasts with several published reports^15,16,43,44^. Average aoPWV in the current study was also slightly higher than what has been reported in the literature; our sample had an average aoPWV of 7.53m/s whereas others have reported 6.8 m/s^45,46^. While there could be a number of contributing factors, it is also theoretically possible that increased stroke volume may register as a higher velocity if arterial remodeling (e.g. diameter or extensibility) has reached its ceiling. According to the principle of continuity in fluid dynamics, if the arterial diameter cannot increase further, then increased stroke volume will result in higher blood velocity^47^. Evidence supports that regular exercise triggers structural adaptations to the heart and the arterial system, lowering resting heart rate^48^ and increasing stroke volume^49^; therefore higher velocities could result from a greater blood volume load verses higher velocities due to arterial stiffening. Future work should therefore take cardiac output, stroke volume, and heart rate into consideration. While outside the scope of the current study, we will be able to interrogate this possibility in our cohort in the future.

This study is not without limitations. Our sample is predominantly white, female, and highly educated. Moreover, the majority of our cohort would be considered highly physically fit. This distribution is not entirely unexpected as the requirement of maximal exercise testing likely biased our results. Lastly, the cross-sectional nature of this report presents interpretive limitations that will be addressed in the future, as we collect additional longitudinal data in this ongoing project. Despite the limitations, strengths are evident in our rigorous approach to measurement. The use of cardiac MRI to assess central arterial stiffness provides more precise measurement of aortic geometry and length thereby increasing reproducibility and reliability of measurements^50^. Furthermore, our newly developed sequence allows for free breathing acquisitions to enable measurement in subjects that cannot hold their breath. 4D flow MRI with radial undersampling captures dynamic changes in blood flow over the cardiac cycle and enables blood flow measurement in all major cerebral arteries^9^.

Lastly, utilization of the VO_2predicted_ measure, calculated from metabolic data collected during an ETT and based on the OUES equation^26^, represents a highly accurate and reliable VO_2_ measure since results are derived directly from individual specific - metabolic gas exchange data providing precise and individualized results.

Overall, our results suggest that average to high CRF or meeting PA recommendations positively modifies the relationship between arterial stiffness and CBF, but that the intensity and type of activity do play a role in the PA-related modifications. Given the evidence supporting a vascular contribution in cognitive decline and AD progression^6,8,9^ and the need to identify modifiable lifestyle factors targeting the preclinical phase of AD, it follows that vascular health should be targeted. While speculative in nature, it appears that the prevention of central arterial aging may attenuate its detrimental impact on cerebral hemodynamics and thereby confer protection from or at least slow the progression of AD-related neuropathological processes or clinical manifestation thereof. Additional research is necessary to further elucidate the exact mechanisms underlying this modifiable and potentially protective risk factor.

## Data Availability

All data produced in the present study are available upon reasonable request to the authors

## 6 CONSENT STATEMENT

Written informed consent was provided by all participants prior to study participation and approval was obtained from the institutional review board.

## 8 AUTHOR CONTRIBUTIONS

BMB, MPG, and OCO designed the experiments. BMB, GSR, TLB, MMJ, LES, AYG, TN, and SRL collected the data. BMB analysed the data, wrote the manuscript and prepared figures. All authors edited, revised, and approved the final version of the manuscript.

## ACKNOWLEDGMENTS

We would like to acknowledge and thank the staff and study participants of the Wisconsin Registry for Alzheimer’s Prevention and the Wisconsin Alzheimer’s Disease Research Center.

## REFERENCES

1. Kim HL, Kim SH. Pulse Wave Velocity in Atherosclerosis. Front Cardiovasc Med [Internet]. Frontiers; 2019 Apr 9 [cited 2024 Apr 22];6. Available from: https://www.frontiersin.org/articles/10.3389/fcvm.2019.00041

2. Zhai FF, Ye YC, Chen SY, Ding FM, Han F, Yang XL, Wang Q, Zhou LX, Ni J, Yao M, Li ML, Jin ZY, Cui LY, Zhang SY, Zhu YC. Arterial Stiffness and Cerebral Small Vessel Disease. Front Neurol. 2018;9:723. PMCID: PMC6121106

3. Haidegger M, Lindenbeck S, Hofer E, Rodler C, Zweiker R, Perl S, Pirpamer L, Kneihsl M, Fandler-Höfler S, Gattringer T, Enzinger C, Schmidt R. Arterial stiffness and its influence on cerebral morphology and cognitive function. Ther Adv Neurol Disord. 2023 Jun 15;16:17562864231180715. PMCID: PMC10285591

4. Li X, Lyu P, Ren Y, An J, Dong Y. Arterial stiffness and cognitive impairment. J Neurol Sci. 2017 Sep 15;380:1–10. PMID: 28870545

5. Jefferson AL, Cambronero FE, Liu D, Moore EE, Neal JE, Terry JG, Nair S, Pechman KR, Rane S, Davis LT, Gifford KA, Hohman TJ, Bell SP, Wang TJ, Beckman JA, Carr JJ. Higher Aortic Stiffness Is Related to Lower Cerebral Blood Flow and Preserved Cerebrovascular Reactivity in Older Adults. Circulation. 2018 Oct 30;138(18):1951–1962. PMCID: PMC6394409

6. Toth P, Tarantini S, Csiszar A, Ungvari Z. Functional vascular contributions to cognitive impairment and dementia: mechanisms and consequences of cerebral autoregulatory dysfunction, endothelial impairment, and neurovascular uncoupling in aging. Am J Physiol Heart Circ Physiol. 2017 Jan 1;312(1):H1–H20. PMCID: PMC5283909

7. Belz GG. Elastic properties and Windkessel function of the human aorta. Cardiovasc Drug Ther. 1995 Feb;9(1):73–83.

8. Zhang H, Wang Y, Lyu D, Li Y, Li W, Wang Q, Li Y, Qin Q, Wang X, Gong M, Jiao H, Liu W, Jia J. Cerebral blood flow in mild cognitive impairment and Alzheimer’s disease: A systematic review and meta-analysis. Ageing Research Reviews. 2021 Nov 1;71:101450.

9. Rivera-Rivera LA, Turski P, Johnson KM, Hoffman C, Berman SE, Kilgas P, Rowley HA, Carlsson CM, Johnson SC, Wieben O. 4D flow MRI for intracranial hemodynamics assessment in Alzheimer’s disease. J Cereb Blood Flow Metab. 2016 Oct;36(10):1718–1730. PMCID: PMC5076787

10. Gaitán JM, Dougherty RJ, Lose S, Maxa KM, Vesperman CJ, Cook DB, Okonkwo OC. Cardiorespiratory fitness bolsters cerebrovascular health in adults at risk for Alzheimer’s disease. Alzheimer’s & Dementia. 2021;17(S10):e049630.

11. Jae SY, Yoon ES, Jung SJ, Jung SG, Park SH, Kim BS, Heffernan KS, Fernhall B. Effect of cardiorespiratory fitness on acute inflammation induced increases in arterial stiffness in older adults. Eur J Appl Physiol. 2013 Aug 1;113(8):2159–2166.

12. Cabral LLP, Freire YA, Browne RAV, Macêdo GAD, Câmara M, Schwade D, Farias- Junior LF, Paulo-Pereira R, Silva RM, Lemos TMAM, Barreira TV, Costa EC. Associations of steps per day and peak cadence with arterial stiffness in older adults. Experimental Gerontology. 2022 Jan 1;157:111628.

13. Liu J, Min L, Liu R, Zhang X, Wu M, Di Q, Ma X. The effect of exercise on cerebral blood flow and executive function among young adults: a double-blinded randomized controlled trial. Sci Rep. Nature Publishing Group; 2023 May 22;13(1):8269.

14. Smith EC, Pizzey FK, Askew CD, Mielke GI, Ainslie PN, Coombes JS, Bailey TG. Effects of cardiorespiratory fitness and exercise training on cerebrovascular blood flow and reactivity: a systematic review with meta-analyses. Am J Physiol Heart Circ Physiol. 2021 Jul 1;321(1):H59–H76. PMID: 34018848

15. Tang A, Eng JJ, Brasher PM, Madden KM, Mohammadi A, Krassioukov AV, Tsang TSM. Physical activity correlates with arterial stiffness in community-dwelling individuals with stroke. J Stroke Cerebrovasc Dis. 2014 Feb;23(2):259–266. PMCID: PMC3828171

16. Gando Y, Murakami H, Kawakami R, Yamamoto K, Kawano H, Tanaka N, Sawada SS, Miyatake N, Miyachi M. Cardiorespiratory Fitness Suppresses Age-Related Arterial Stiffening in Healthy Adults: A 2-Year Longitudinal Observational Study. J Clin Hypertens (Greenwich). 2016 Apr;18(4):292–298. PMCID: PMC8031982

17. Park SY, Kwak YS, Pekas EJ. Impacts of aquatic walking on arterial stiffness, exercise tolerance, and physical function in patients with peripheral artery disease: a randomized clinical trial. J Appl Physiol (1985). 2019 Oct 1;127(4):940–949. PMID: 31369328

18. Tomoto T, Verma A, Kostroske K, Tarumi T, Patel NR, Pasha EP, Riley J, Tinajero CD, Hynan LS, Rodrigue KM, Kennedy KM, Park DC, Zhang R. One-year aerobic exercise increases cerebral blood flow in cognitively normal older adults. J Cereb Blood Flow Metab. 2023 Mar;43(3):404–418. PMCID: PMC9941859

19. Tomoto T, Liu J, Tseng BY, Pasha EP, Cardim D, Tarumi T, Hynan LS, Munro Cullum C, Zhang R. One-Year Aerobic Exercise Reduced Carotid Arterial Stiffness and Increased Cerebral Blood Flow in Amnestic Mild Cognitive Impairment. J Alzheimers Dis. 2021;80(2):841–853. PMID: 33579857

20. Dougherty RJ, Boots EA, Lindheimer JB, Stegner AJ, Van Riper S, Edwards DF, Gallagher CL, Carlsson CM, Rowley HA, Bendlin BB, Asthana S, Hermann BP, Sager MA, Johnson SC, Okonkwo OC, Cook DB. Fitness, independent of physical activity is associated with cerebral blood flow in adults at risk for Alzheimer’s disease. Brain Imaging Behav. 2020 Aug;14(4):1154–1163. PMCID: PMC6733668

21. Franklin BA, Eijsvogels TMH, Pandey A, Quindry J, Toth PP. Physical activity, cardiorespiratory fitness, and cardiovascular health: A clinical practice statement of the ASPC Part I: Bioenergetics, contemporary physical activity recommendations, benefits, risks, extreme exercise regimens, potential maladaptations. Am J Prev Cardiol. 2022 Oct 13;12:100424. PMCID: PMC9586848

22. Myers J, Kaykha A, George S, Abella J, Zaheer N, Lear S, Yamazaki T, Froelicher V. Fitness versus physical activity patterns in predicting mortality in men. The American Journal of Medicine. 2004 Dec 15;117(12):912–918.

23. Miller KR, McClave SA, Jampolis MB, Hurt RT, Krueger K, Landes S, Collier B. The Health Benefits of Exercise and Physical Activity. Curr Nutr Rep. 2016 Sep 1;5(3):204–212.

24. Johnson SC, Koscik RL, Jonaitis EM, Clark LR, Mueller KD, Berman SE, Bendlin BB, Engelman CD, Okonkwo OC, Hogan KJ, Asthana S, Carlsson CM, Hermann BP, Sager MA. The Wisconsin Registry for Alzheimer’s Prevention: A review of findings and current directions. Alzheimers Dement (Amst). 2018;10:130–142. PMCID: PMC5755749

25. Dougherty RJ, Jonaitis EM, Gaitán JM, Lose SR, Mergen BM, Johnson SC, Okonkwo OC, Cook DB. Cardiorespiratory fitness mitigates brain atrophy and cognitive decline in adults at risk for Alzheimer’s disease. Alzheimers Dement (Amst). 2021;13(1):e12212. PMCID: PMC8274307

26. Dougherty RJ, Lindheimer JB, Stegner AJ, Van Riper S, Okonkwo OC, Cook DB. An Objective Method to Accurately Measure Cardiorespiratory Fitness in Older Adults Who Cannot Satisfy Widely Used Oxygen Consumption Criteria. J Alzheimers Dis. 2018;61(2):601–611. PMCID: PMC5745283

27. Kaminsky LA, Arena R, Myers J. Reference Standards for Cardiorespiratory Fitness Measured With Cardiopulmonary Exercise Testing. Mayo Clinic Proceedings. 2015 Nov;90(11):1515–1523.

28. Stewart AL, Mills KM, King AC, Haskell WL, Gillis D, Ritter PL. CHAMPS physical activity questionnaire for older adults: outcomes for interventions. Med Sci Sports Exerc. 2001 Jul;33(7):1126–1141. PMID: 11445760

29. Global Recommendations on Physical Activity for Health [Internet]. Geneva: World Health Organization; 2010 [cited 2024 Oct 2]. Available from: http://www.ncbi.nlm.nih.gov/books/NBK305057/ PMID: 26180873

30. Grant S. Roberts, Kevin M. Johnson, Steven R. Keskemeti, Ozioma Okonkwo, Sarah Lose, Laura Eisenmenger, Oliver Wieben. Feasibility of a Free-Breathing 2D Phase Contrast Sequence for Aortic Pulse Wave Velocity Measurements. ISMRM & SMRT Virtual Conference & Exhibition Abstract # 2262 [Internet]. 2023 [cited 2024 Jan 31]; Available from: https://cds.ismrm.org/protected/20MProceedings/PDFfiles/2262.html

31. Dogui A, Redheuil A, Lefort M, DeCesare A, Kachenoura N, Herment A, Mousseaux E. Measurement of aortic arch pulse wave velocity in cardiovascular MR: Comparison of transit time estimators and description of a new approach. Journal of Magnetic Resonance Imaging. 2011;33(6):1321–1329.

32. Gu T, Korosec FR, Block WF, Fain SB, Turk Q, Lum D, Zhou Y, Grist TM, Haughton V, Mistretta CA. PC VIPR: a high-speed 3D phase-contrast method for flow quantification and high-resolution angiography. AJNR Am J Neuroradiol. 2005 Apr;26(4):743–749. PMCID: PMC7977085

33. Johnson KM, Lum DP, Turski PA, Block WF, Mistretta CA, Wieben O. Improved 3D phase contrast MRI with off-resonance corrected dual echo VIPR. Magn Reson Med. 2008 Dec;60(6):1329–1336. PMCID: PMC2778052

34. Maxa KM, Hoffman C, Rivera-Rivera LA, Motovylyak A, Turski PA, Mitchell CKC, Ma Y, Berman SE, Gallagher CL, Bendlin BB, Asthana S, Sager MA, Hermann BP, Johnson SC, Cook DB, Wieben O, Okonkwo OC. Cardiorespiratory Fitness Associates with Cerebral Vessel Pulsatility in a Cohort Enriched with Risk for Alzheimer’s Disease. Brain Plast. 2020 Oct 1;5(2):175–184. PMCID: PMC7685671

35. Sabra D, Intzandt B, Desjardins-Crepeau L, Langeard A, Steele CJ, Frouin F, Hoge RD, Bherer L, Gauthier CJ. Sex moderations in the relationship between aortic stiffness, cognition, and cerebrovascular reactivity in healthy older adults. PLoS One. 2021 Sep 28;16(9):e0257815. PMCID: PMC8478243

36. Fico BG, Miller KB, Rivera-Rivera LA, Corkery AT, Pearson AG, Eisenmann NA, Howery AJ, Rowley HA, Johnson KM, Johnson SC, Wieben O, Barnes JN. The Impact of Aging on the Association Between Aortic Stiffness and Cerebral Pulsatility Index. Front Cardiovasc Med. 2022 Feb 9;9:821151. PMCID: PMC8863930

37. DuBose LE, Boles Ponto LL, Moser DJ, Harlynn E, Reierson L, Pierce GL. Higher Aortic Stiffness is Associated with Lower Global Cerebrovascular Reserve Among Older Humans. Hypertension. 2018 Aug;72(2):476–482. PMCID: PMC6261448

38. Van Bortel LM, Laurent S, Boutouyrie P, Chowienczyk P, Cruickshank JK, De Backer T, Filipovsky J, Huybrechts S, Mattace-Raso FUS, Protogerou AD, Schillaci G, Segers P, Vermeersch S, Weber T, Artery Society, European Society of Hypertension Working Group on Vascular Structure and Function, European Network for Noninvasive Investigation of Large Arteries. Expert consensus document on the measurement of aortic stiffness in daily practice using carotid-femoral pulse wave velocity. J Hypertens. 2012 Mar;30(3):445–448. PMID: 22278144

39. Nobili F, Rodriguez G, Marenco S, De Carli F, Gambaro M, Castello C, Pontremoli R, Rosadini G. Regional cerebral blood flow in chronic hypertension. A correlative study. Stroke. 1993 Aug;24(8):1148–1153. PMID: 8342188

40. Lipsitz LA, Mukai S, Hamner J, Gagnon M, Babikian V. Dynamic regulation of middle cerebral artery blood flow velocity in aging and hypertension. Stroke. 2000 Aug;31(8):1897–1903. PMID: 10926954

41. Lipsitz LA, Gagnon M, Vyas M, Iloputaife I, Kiely DK, Sorond F, Serrador J, Cheng DM, Babikian V, Cupples LA. Antihypertensive Therapy Increases Cerebral Blood Flow and Carotid Distensibility in Hypertensive Elderly Subjects. Hypertension. American Heart Association; 2005 Feb;45(2):216–221.

42. Tarumi T, Gonzales MM, Fallow B, Nualnim N, Pyron M, Tanaka H, Haley AP. Central artery stiffness, neuropsychological function, and cerebral perfusion in sedentary and endurance-trained middle-aged adults. Journal of Hypertension. 2013 Dec;31(12):2400.

43. Tanaka H, DeSouza CA, Seals DR. Absence of age-related increase in central arterial stiffness in physically active women. Arterioscler Thromb Vasc Biol. 1998 Jan;18(1):127–132. PMID: 9445266

44. Tanaka H, Dinenno FA, Monahan KD, Clevenger CM, DeSouza CA, Seals DR. Aging, Habitual Exercise, and Dynamic Arterial Compliance. Circulation. American Heart Association; 2000 Sep 12;102(11):1270–1275.

45. van Hout MJ, Dekkers IA, Westenberg JJ, Schalij MJ, Widya RL, de Mutsert R, Rosendaal FR, de Roos A, Jukema JW, Scholte AJ, Lamb HJ. Normal and reference values for cardiovascular magnetic resonance-based pulse wave velocity in the middle-aged general population. Journal of Cardiovascular Magnetic Resonance. 2021 Apr 19;23(1):46.

46. Parikh JD, Hollingsworth KG, Kunadian V, Blamire A, MacGowan GA. Measurement of pulse wave velocity in normal ageing: comparison of Vicorder and magnetic resonance phase contrast imaging. BMC Cardiovascular Disorders. 2016 Feb 19;16(1):50.

47. Menon K, Hu Z, Marsden AL. Cardiovascular fluid dynamics: a journey through our circulation. Flow. 2024 Jan;4:E7.

48. Rosenwinkel ET, Bloomfield DM, Arwady MA, Goldsmith RL. Exercise and autonomic function in health and cardiovascular disease. Cardiol Clin. 2001 Aug;19(3):369–387. PMID: 11570111

49. Hellsten Y, Nyberg M. Cardiovascular Adaptations to Exercise Training. Compr Physiol. 2015 Dec 15;6(1):1–32. PMID: 26756625

50. Houriez--Gombaud-Saintonge S, Mousseaux E, Bargiotas I, De Cesare A, Dietenbeck T, Bouaou K, Redheuil A, Soulat G, Giron A, Gencer U, Craiem D, Messas E, Bollache E, Chenoune Y, Kachenoura N. Comparison of different methods for the estimation of aortic pulse wave velocity from 4D flow cardiovascular magnetic resonance. Journal of Cardiovascular Magnetic Resonance. 2019 Dec 12;21(1):75.

