## Supplemental Figure 1 for "Cardiorespiratory Fitness Modifies the Relationship Between Arterial Stiffness and Cerebral Blood Flow Independent of Physical Activity"

### sFigure 1 | The relationship between arterial stiffness and CBF is attenuated by fitness level.

The figure depicts the results from covariate adjusted, linear regression models testing the statistical moderation of fitness on the relationship between aoPWV and CBF; observed data points are plotted whereas regression lines and confidence intervals are covariate adjusted based on mean values from the model. Covariates included age, sex, WHR, HTN, APOE ε4+, and PA Rec Met. **Average and high fitness attenuated the adverse relationship between arterial stiffness and CBF for (A) Total and (B) Global flow and in the (C) ICA, (D) Cav. ICA and (E) MCA.**

*Abbreviations:* ***APOE ε4+*** *= APOE ε4 allele carriage;* ***aoPWV*** *= aortic pulse wave velocity;* ***BA*** *= Basilar artery;* ***Cav. ICA*** *= cavernous ICA;* ***Fitness*** *= Low, average, and high cardiorespiratory fitness groups were defined based on their calculated* $VO_{2predicted}$ *falling within the fitness percentile ranges for age and sex: ≤49%, 50%-60%, and ≥ 70%, respectively;* ***Global Q*** *= global flow; the average of seven cerebral arteries;* ***HTN*** *= hypertension;* ***ICA*** *= internal carotid arteries;* ***MCA*** *= middle cerebral artery;* ***PA Rec Met*** *= Physical Activity Recommendations of 150 minutes of moderate-intensity activity were met (yes/no);* ***Total Q*** *= total flow; the sum of the ICAs and the BA;* ***WHR*** *= waist to hip ratio.*

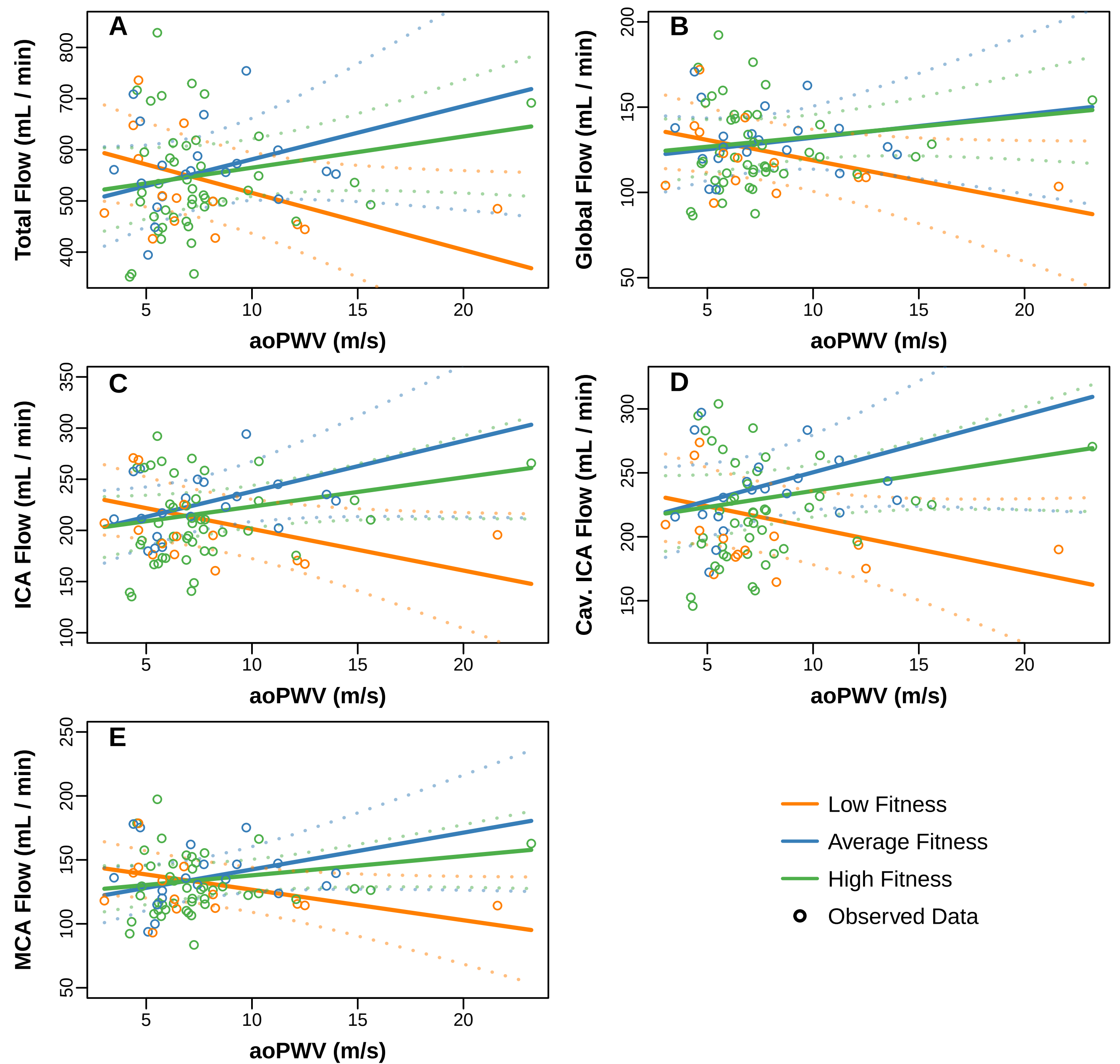

A
B
C
D
E
Total Flow (mL / min)
Global Flow (mL / min)
ICA Flow (mL / min)
Cav. ICA Flow (mL / min)
MCA Flow (mL / min)
aoPWV (m/s)
Low Fitness
Average Fitness
High Fitness
Observed Data
