## Supplemental Figure 2 for "Cardiorespiratory Fitness Modifies the Relationship Between Arterial Stiffness and Cerebral Blood Flow Independent of Physical Activity"

**sFigure 2 | The relationship between arterial stiffness and CBF is attenuated by meeting physical activity recommendations.**

The figure depicts the results from covariate adjusted, linear regression models testing the statistical moderation of physical activity (PA Rec Met) on the relationship between aoPWV and CBF; observed data points are plotted whereas regression lines and confidence intervals are covariate adjusted based on mean values from the model. Covariates included age, sex, WHR, HTN, APOE ε4+,and $VO_{2predicted}$. **Meeting the physical activity recommendations (PA Rec Met) attenuated the adverse relationship between arterial stiffness and CBF for (A) Total and (B) Global flow and in (E) the MCA.**

*Abbreviations:* ***APOE ε4****+ = APOE ε4 allele carriage;* ***aoPWV*** *= aortic pulse wave velocity;* ***BA =*** *Basilar artery;* ***Cav. ICA*** *= cavernous ICA;* ***Global Q*** *= global flow; the average of seven cerebral arteries;* ***HTN*** *= hypertension;* ***ICA*** *= internal carotid arteries;* ***MCA*** *= middle cerebral artery;* ***PA Rec Met*** *= Physical Activity Recommendations of 150 minutes of moderate-intensity activity were met (yes/no);* ***Total Q*** *= total flow; the sum of the ICAs and the BA;* $VO_{2predicted}$ *= predicted maximal oxygen consumption based on the oxygen uptake efficiency slope;* ***WHR*** *= waist to hip ratio.*

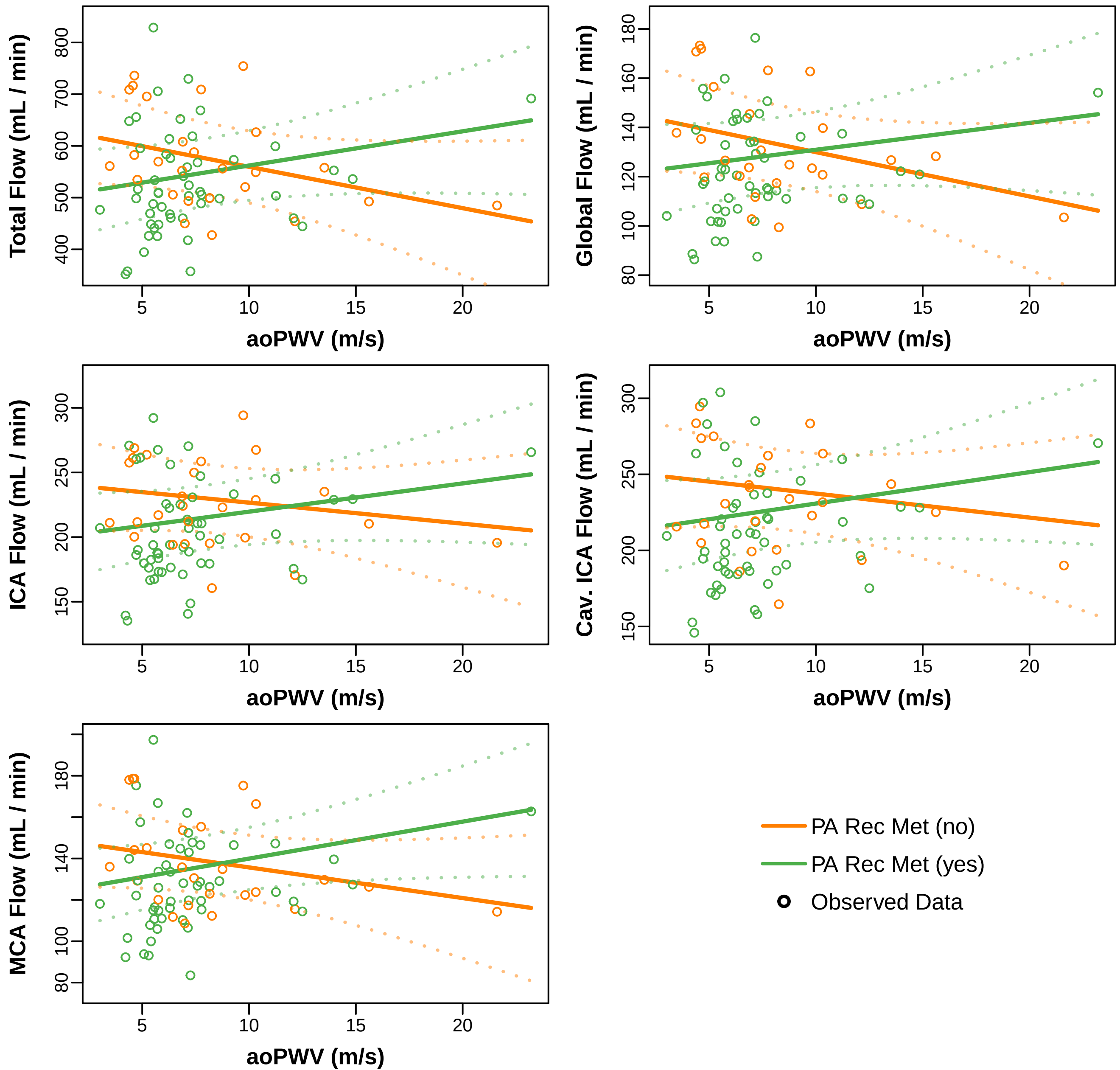

Total Flow (mL / min)
Global Flow (mL / min)
ICA Flow (mL / min)
Cav. ICA Flow (mL / min)
MCA Flow (mL / min)
aoPWV (m/s)
PA Rec Met (no)
PA Rec Met (yes)
Observed Data
